# Shared host-genetic architecture between gut microbiota and internalizing psychopathology

**DOI:** 10.64898/2026.05.31.26354553

**Authors:** Pedro Velez-Pardo, Roger Jose Solano, Adriana Maria Quinchia-Figueroa, Ricardo Montoya Monsalve, Nadia Semenova Moratto-Vasquez

**Affiliations:** Universidad EIA, Km 2 + 200, Alto de Las Palmas, Envigado, Colombia; Universidad CES, Calle 10A # 22-04, Medellin, Colombia

**Keywords:** Mendelian randomization, Gut microbiome, Internalizing psychopathology, Depression, Neuroticism, Insomnia, Genomic SEM

## Abstract

Whether gut microbial composition is causally linked to mental illness, or merely correlated with it, remains unresolved. Using genetic variants as natural instruments (Mendelian randomization, MR), we tested the genetically predicted effects of 211 gut microbial taxa on nine psychiatric and psychopathology-related phenotypes, using the largest available genome-wide association studies. Across 1,898 valid tests, seven taxon-outcome associations passed false-discovery-rate correction (FDR < 0.05), and all fell on the internalizing spectrum (depression, neuroticism and insomnia) rather than on bipolar disorder or schizophrenia; they included a protective association of the Mollicutes/Tenericutes clade with depression (β = -0.073, p = 1.5 ×10^-6^) and of Butyrivibrio with neuroticism, and a deleterious association of Betaproteobacteria with neuroticism. Conservative tests tempered any per-locus causal reading: Bayesian colocalization gave a posterior probability of a shared causal variant (PP.H4) < 0.05 at every locus, and a summary-data causal test (CAUSE) found 0 of 45 taxon-outcome pairs genuinely causal; yet the direction of effect matched the protective-versus-deleterious hypothesis in 33 of 45 pairs (binomial p = 1.2 ×10^-3^). Modelling the shared genetics of the nine phenotypes placed these taxa specifically on a latent internalizing factor (correlation 0.48 with a separate psychotic factor) and, in a bifactor model, on internalizing-specific genetic variance beyond a general psychopathology factor. Selected gut microbial taxa and internalizing psychopathology therefore appear to share host genetics rather than a direct microbe-to-disorder causal chain. We release the full analysis as an open resource for larger microbiome studies, brain-tissue follow-up and experimental tests of candidate mechanisms.

## Introduction

Psychiatric disorders are among the leading causes of disability worldwide, affecting more than one in eight people during their lifetime and accounting for an estimated 16% of global years lived with disability (GBD 2019 Mental Disorders Collaborators, 2022; World Health Organization, 2022). The molecular mechanisms of common psychiatric conditions remain only partially understood, and identifying modifiable biological contributors to psychiatric risk is a continuing priority.

The gut microbiome has emerged as a candidate axis through which peripheral biology may influence brain function and behaviour (Cryan et al., 2019; Mayer et al., 2022). Work in rodents and humans documents bidirectional communication between commensal bacteria and the central nervous system via short-chain fatty acid (SCFA) signalling, tryptophan and kynurenine metabolism, regulation of systemic inflammation through interleukin-6 (IL-6) and C-reactive protein (CRP), γ-aminobutyric acid (GABA) neurotransmission, and vagal afferent activation (Dalile et al., 2019; Strandwitz et al., 2019). Observational studies report altered microbial composition in major depressive disorder (MDD), bipolar disorder (BD), schizophrenia and anxiety disorders, with recurrent depletion of butyrate-producing taxa (Amin et al., 2023; Jiang et al., 2015; Valles-Colomer et al., 2019). However, observational designs cannot easily separate cause from consequence: illness, medication and lifestyle all reshape the microbiome, opening the door to reverse causation and confounding (Vujkovic-Cvijin et al., 2020).

Mendelian randomization (MR) addresses causation using germline genetic variants as instruments for a putative exposure, approximating a natural randomized experiment (Davey Smith and Hemani, 2014; Sanderson et al., 2022). The MiBioGen consortium meta-analysis, which identified host-genetic determinants of 211 microbial taxa across 18,340 European individuals by 16S rRNA sequencing of the V4 region, made it possible to instrument gut microbiome composition at scale (Kurilshikov et al., 2021).

A growing number of MR studies have interrogated taxon-outcome relationships in psychiatry, including disorder-specific reports on MDD (Chen et al., 2022), BD (Zhao et al., 2025), schizophrenia (Zhou et al., 2024), insomnia (Yang et al., 2023), anxiety (Li et al., 2024) and ADHD (Wang et al., 2023), and broader scans across multiple phenotypes (Ni et al., 2022; Wu et al., 2025; Xiao et al., 2023) or integrating brain-structural endophenotypes (Ye et al., 2025). Three limitations persist. First, most studies analyse one or a few outcomes at a time, leaving open whether microbial signals are disorder-specific or instead concentrate on shared latent psychiatric architecture, a question central to debates about the dimensional structure of psychiatric nosology (Caspi and Moffitt, 2018; Grotzinger et al., 2022; Lee et al., 2019). Second, many discovery GWAS used as outcomes have been superseded by substantially larger releases since 2023, including PGC PTSD Freeze 3 (Nievergelt et al., 2024), PGC obsessive-compulsive disorder (OCD) (Strom et al., 2025), the PGC anxiety meta-analysis (Strom et al., 2026), the MDD meta-analysis (Adams et al., 2025) and PGC3 schizophrenia (Trubetskoy et al., 2022). Third, prior work has rarely combined univariable MR with multivariable cross-disorder analyses, formal mediation, and the most conservative tests (Bayesian colocalization and CAUSE for correlated horizontal pleiotropy), leaving room for selective reporting in a high-dimensional analytic space.

Here we combine, within a single analytic pipeline applied uniformly to nine psychiatric and psychopathology-related outcomes: the most recent psychiatric GWAS available as of 2026; univariable and multivariable MR; two-step mediation through tryptophan, kynurenine, CRP and IL-6; LDSC; Bayesian colocalization; MAGMA; CAUSE; MR-BMA with a permutation null; a two-factor Genomic SEM; and exploratory cis-eQTL prioritization. We report the full atlas as a resource, but our principal finding is focused: the directional MR signal concentrates on the internalizing domain, and selected taxa share host-genetic architecture specifically with the internalizing dimension of psychopathology rather than exerting direct per-locus causation.

## Methods

### Study design

We performed a two-sample MR study of the genetically instrumented effects of gut microbial composition on nine psychiatric and psychopathology-related outcomes, with mediation through four candidate biological mediators. The protocol was pre-registered on the Open Science Framework before data analysis (lock date 17 May 2026; DOI 10.17605/OSF.IO/FVGQ2); 25 amendments are declared transparently across four revisions (full log in Supplementary Materials and on OSF). Reporting follows STROBE-MR (Skrivankova et al., 2021). We use ’genetically instrumented associations’ rather than ’causal effects’ for taxon-outcome relationships, reserving causal language for the conservative CAUSE framework. The ten-layer framework is summarized in Figure 1.

**Figure 1.**
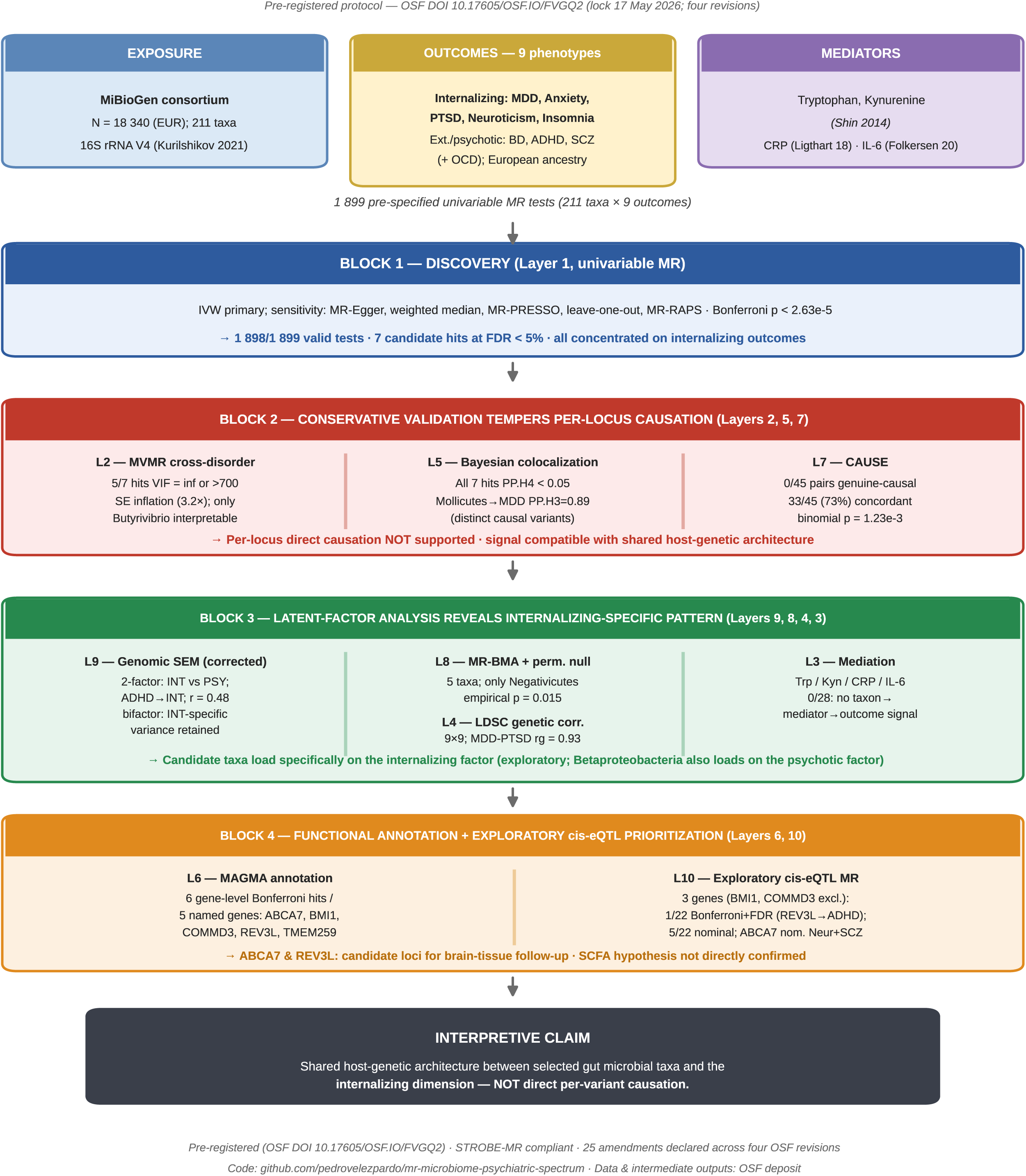
Study design and ten-layer analytical framework. Overview of the two-sample Mendelian randomization design (211 MiBioGen taxa × 9 psychiatric and psychopathology-related outcomes, plus 4 candidate mediators) and the ten analytical layers, organized as a discovery to conservative-validation to latent-factor to exploration cascade. The OSF pre-registration and amendment log are indicated; the principal interpretive claim (shared host-genetic architecture with the internalizing dimension) is shown in the final panel.

### Exposures, outcomes and mediators

Genetic instruments for 211 MiBioGen taxa (9 phyla, 16 classes, 20 orders, 35 families, 131 genera) were obtained from the MiBioGen consortium (Kurilshikov et al., 2021); effect sizes are per standard-deviation increase in log-transformed relative abundance. Nine outcomes spanned internalizing, neurodevelopmental and psychotic domains: MDD (no23andMe/noUKBB; 357,636 cases / 1,281,936 controls; Adams et al., 2025), anxiety (122,341 / 729,881; Strom et al., 2026), BD (noUKBB; 40,463 / 313,436; Mullins et al., 2021), insomnia (UK Biobank, ukb-b-3957; N = 462,341; Elsworth et al., 2020), neuroticism (UK Biobank, ukb-b-4630; N = 374,323; Elsworth et al., 2020), ADHD (38,691 / 186,843; Demontis et al., 2023), PTSD (PGC Freeze 3; 137,136 / 1,085,746; Nievergelt et al., 2024), schizophrenia (PGC3; 53,386 / 77,258; Trubetskoy et al., 2022) and OCD (22,717 / 988,884; Strom et al., 2025). We note that neuroticism and insomnia are trait/symptom rather than diagnostic phenotypes, and that the panel does not cover the full DSM-5 nosology. Four mediators were pre-specified: tryptophan and kynurenine (Shin et al., 2014), CRP (Ligthart et al., 2018) and IL-6 (Folkersen et al., 2020). Accessions, sample sizes and download details are in Supplementary Table 1.

### Instrument selection and analytical layers

For each taxon, SNPs at p < 1 ×10^-5^ were LD-clumped at r^2^ < 0.001 within 10 Mb (1000 Genomes EUR), excluding palindromic SNPs at intermediate frequency. Across the 211 taxa, mean F per taxon ranged 20.3-25.9 (median 21.6; minimum per-SNP F > 19); the narrow range reflects the p < 1 ×10^-5^ threshold and is addressed in Limitations as a weak-instrument concern. Per-taxon F-statistics are in Supplementary Table 13, the instrument-selection flow in Supplementary Figure S1 and the F-statistic distribution in Supplementary Figure S3. Harmonization used TwoSampleMR (action = 2) (Hemani et al., 2018) in GRCh37.

The ten layers were: (1) univariable MR (random-effects IVW primary, with MR-Egger (Bowden et al., 2015), weighted median (Bowden et al., 2016), MR-PRESSO (Verbanck et al., 2018), leave-one-out and MR-RAPS (Zhao et al., 2020); Bonferroni p < 2.63 ×10^-5^ = 0.05/1,899); (2) multivariable cross-disorder MR, with conditional F and variance-inflation diagnostics, restricted to a taxon-only instrument set after diagnosing severe standard-error inflation in the union-of-instruments specification (Sanderson et al., 2021); (3) two-step mediation (10,000 bootstrap CIs); (4) LDSC (Bulik-Sullivan et al., 2015); (5) Bayesian colocalization (coloc.abf, ±500 kb) (Giambartolomei et al., 2014), with multiple-causal-variant methods (coloc-SuSiE) noted as a constraint (Wallace, 2021); (6) MAGMA v1.10 (Bonferroni p < 2.72 ×10^-6^ = 0.05/18,415 genes) (de Leeuw et al., 2015); (7) CAUSE (Morrison et al., 2020); (8) MR-BMA with a 200-iteration permutation null (Zuber et al., 2020); (9) Genomic SEM (Grotzinger et al., 2019), comparing one-factor, two-factor and bifactor structures (see below); and (10) exploratory cis-eQTL gene prioritization using eQTLGen whole blood (Vosa et al., 2021) with DGIdb v5 annotation (Cannon et al., 2024). Analyses used R 4.x; code and intermediate outputs are on the project repository and OSF.

### Latent-factor analysis

Genomic SEM (Grotzinger et al., 2019) was fitted to the LDSC genetic covariance matrix of eight outcomes (OCD excluded for insufficient instruments) using diagonally weighted least squares. Our pre-registered protocol specified a two-factor model assigning ADHD to the externalizing/psychotic factor. Because ADHD correlates genetically with the internalizing block far more strongly (rg with MDD 0.61, PTSD 0.68) than with bipolar disorder or schizophrenia (0.22 and 0.20), and because the pre-registered assignment produced an improper solution (a Heywood case with an inflated inter-factor correlation), we assigned ADHD to the internalizing/distress factor in the primary model; this is a genetic-architecture grouping, distinct from ADHD’s clinical classification as a neurodevelopmental disorder. We additionally fitted a bifactor model with a general psychopathology factor and an orthogonal internalizing-specific factor. This data-driven deviation from the pre-registered factor structure is documented as a declared amendment on OSF. Taxon-to-factor effects were estimated by inverse-variance-weighted projection of the per-outcome univariable estimates onto the factor loadings.

### Use of AI-assisted tools in the analysis

A large language model-based AI assistant (Claude, Anthropic) was used to reproduce and cross-check the corrected Genomic SEM re-analysis and to generate plotting code for the display items from the study’s own computed statistics. The primary Mendelian randomization pipeline was specified and executed by the authors. All code, analyses and results were reviewed and verified by the authors, who take full responsibility for the content. No generative AI was used to create, alter or fabricate data, results or figures.

## Results

### Discovery: seven candidate taxa concentrate on internalizing outcomes

Of 1,899 attempted univariable tests (211 taxa × 9 outcomes), 1,898 returned valid IVW estimates. Two redundant taxonomic entries representing one clade (class Mollicutes and phylum Tenericutes) reached Bonferroni-stringent significance with a protective association on MDD (β = -0.073 per SD increase in log-abundance, SE = 0.015, p = 1.45 ×10^-6^; FDR = 0.0014; MR-PRESSO global p = 0.85). Five further pairs reached Benjamini-Hochberg FDR < 5%: the Negativicutes/Selenomonadales clade on insomnia (β = +0.034, p = 4.28 ×10^-5^, deleterious); class Betaproteobacteria on neuroticism (β = +0.200, p = 8.58 ×10^-5^, deleterious); genus Butyrivibrio on neuroticism (β = -0.086, p = 9.30 ×10^-5^, protective); and genus Tyzzerella3 on neuroticism (β = -0.084, p = 1.01 ×10^-4^, protective). After de-duplication this corresponds to seven entries across five biological clades (Figure 2, Table 1).

**Figure 2.**
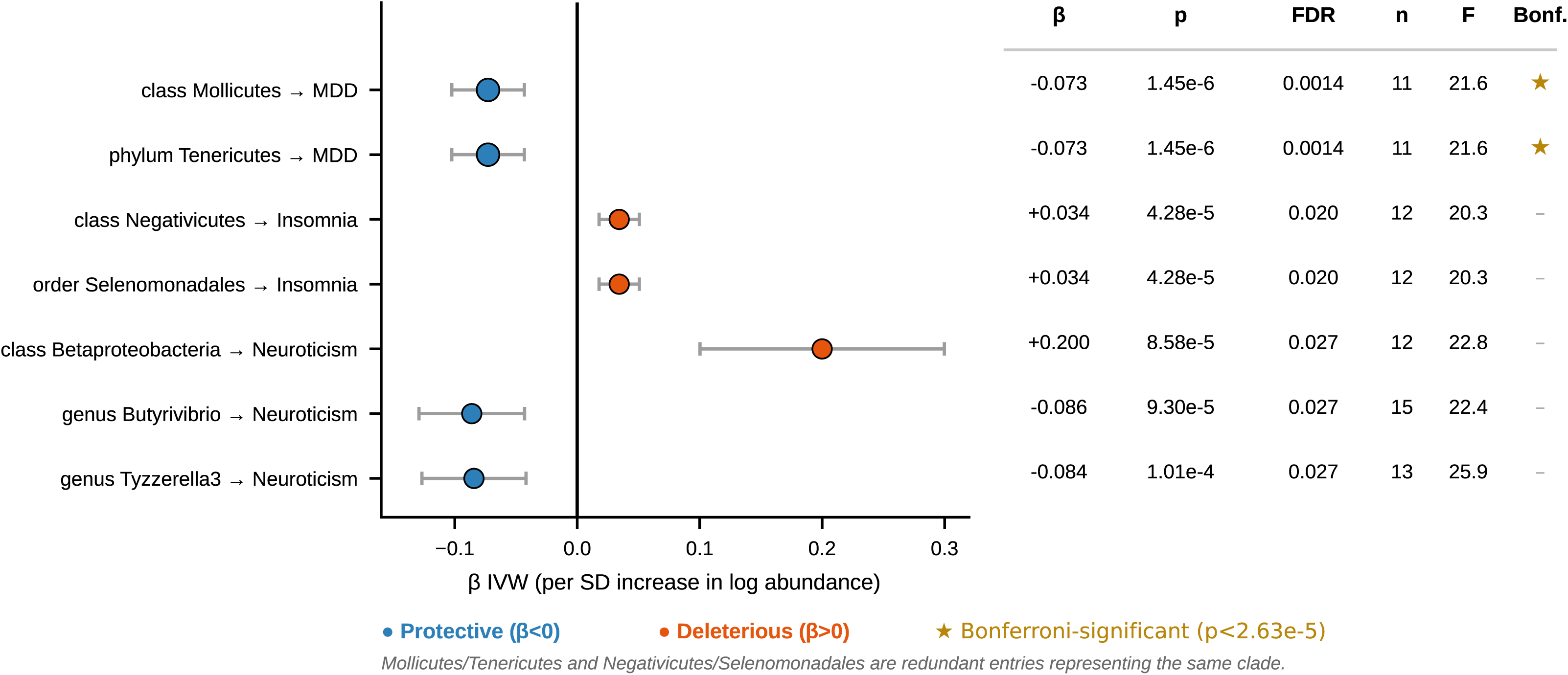
Univariable MR discovery: seven candidate taxon-outcome hits. Forest plot of inverse-variance-weighted (IVW) β with 95% confidence interval for the seven taxon-outcome pairs reaching FDR < 0.05, with FDR, Bonferroni status, number of SNPs and mean F. Hits are coloured by direction (protective β < 0, blue; deleterious β > 0, orange). All seven concentrate on internalizing outcomes (MDD, neuroticism, insomnia). Mollicutes/Tenericutes and Negativicutes/Selenomonadales are redundant taxonomic entries representing the same clades. The full 211 × 9 landscape is provided as Supplementary Figure S2 and as a volcano plot in Supplementary Figure S8.

**Table 1.** Layer 1 candidate hits: comprehensive sensitivity panel. For each of the seven candidate pairs: taxon, outcome, hypothesized direction, number of SNPs, mean F, IVW β/SE/p, FDR (Benjamini-Hochberg), Bonferroni status, MR-Egger and weighted-median estimates and p-values, MR-Egger intercept p, Cochran Q p, Bayesian colocalization PP.H4 and dominant posterior, and CAUSE conclusion.

| Hit # | Taxon | Outcome | Direction (hypothesis) | n SNPs | mean F | $\beta$ IVW | SE IVW | p IVW | FDR-BH | Bonferroni (0.05/1,899) | $\beta$ MR-Egger | p MR-Egger | $\beta$ Weighted Median | p Weighted Median | Egger intercept p | Cochran Q p (IVW) | Coloc PP.H4 | Coloc dominant | CAUSE conclusion |
| --- | --- | --- | --- | --- | --- | --- | --- | --- | --- | --- | --- | --- | --- | --- | --- | --- | --- | --- | --- |
| 1 | c. Mollicutes | MDD | Protective | 11 | 21.6 | -0.07277 | 0.0151 | 1.45e-06 | 0.00138 | Yes | -0.0132 | 0.784 | -0.0696 | 0.000801 | 0.211 | 0.814 | 0.00128 | distinct causal variants (PP.H3 ≥ 0.50) | NO_EFFECT |
| 2 | p. Tenericutes | MDD | Protective | 11 | 21.6 | -0.07277 | 0.0151 | 1.45e-06 | 0.00138 | Yes | -0.0132 | 0.784 | -0.0696 | 0.00102 | 0.211 | 0.814 | 0.00128 | distinct causal variants (PP.H3 ≥ 0.50) | NO_EFFECT |
| 3 | c. Negativicutes | Insomnia | Deleterious | 12 | 20.3 | 0.03427 | 0.00837 | 4.28e-05 | 0.0203 | No | 0.0449 | 0.129 | 0.0443 | 0.000213 | 0.688 | 0.457 | 0.00289 | exposure-only association (PP.H1 ≥ 0.50) | NO_EFFECT |
| 4 | o. Selenomonadales | Insomnia | Deleterious | 12 | 20.3 | 0.03427 | 0.00837 | 4.28e-05 | 0.0203 | No | 0.0449 | 0.129 | 0.0443 | 0.000184 | 0.688 | 0.457 | 0.00289 | exposure-only association (PP.H1 ≥ 0.50) | NO_EFFECT |
| 5 | c. Betaproteobacteria | Neuroticism | Deleterious | 12 | 22.8 | 0.2 | 0.0509 | 8.58e-05 | 0.0273 | No | -0.098 | 0.553 | 0.162 | 0.00766 | 0.0802 | 0.127 | 0.0173 | exposure-only association (PP.H1 ≥ 0.50) | NO_EFFECT |
| 6 | g. Butyrivibrio | Neuroticism | Protective | 15 | 22.4 | -0.08605 | 0.022 | 9.3e-05 | 0.0273 | No | -0.00368 | 0.971 | -0.0748 | 0.00298 | 0.412 | 0.0527 | 0.00518 | exposure-only association (PP.H1 ≥ 0.50) | NO_EFFECT |
| 7 | g. Tyzzerella3 | Neuroticism | Protective | 13 | 25.9 | -0.08429 | 0.0217 | 0.000101 | 0.0273 | No | -0.148 | 0.257 | -0.0909 | 0.00262 | 0.611 | 0.56 | 0.00277 | distinct causal variants (PP.H3 ≥ 0.50) | NO_EFFECT |

Critically, every discovery hit fell within the internalizing domain (MDD, neuroticism, insomnia); none reached FDR for ADHD, PTSD, schizophrenia, BD, anxiety or OCD. Direction was concordant across IVW and weighted median for 7/7 pairs and across MR-Egger slope for 6/7 (the exception, Betaproteobacteria on neuroticism, showed an imprecise sign-discordant Egger slope, p = 0.55, with a non-significant intercept). Cochran’s Q was non-significant for all hits (full sensitivity, heterogeneity and MR-Egger intercept estimates in Supplementary Tables 10-12; a focused per-hit forest plot in Supplementary Figure S9; the full 211 × 9 p-value landscape in Supplementary Figure S2 and a volcano plot in Supplementary Figure S8). This internalizing concentration motivated the conservative validation and latent-factor analyses below.

### Conservative validation tempers the per-locus causal interpretation

Three conservative tests modified the causal interpretation. First, multivariable cross-disorder MR restricted to taxon-only instruments was structurally over-parameterized for five of seven entries (variance-inflation factor infinite or > 700 for an eight-exposure model instrumented by 6-14 taxon SNPs); only Butyrivibrio (32 taxon-only instruments) was numerically interpretable, retaining within-model Bonferroni-significant negative effects on MDD (β = -0.043, p = 3.0 ×10^-5^) and neuroticism (β = -0.082, p = 1.6 ×10^-6^). The originally striking Mollicutes/Tenericutes-MDD signal did not survive this specification (p = 0.11; full multivariable-MR estimates and variance-inflation diagnostics in Supplementary Tables 14 and 15). Second, Bayesian colocalization returned PP.H4 < 0.05 at every candidate locus; the Mollicutes/Tenericutes and Tyzzerella3 loci showed PP.H3 dominance (distinct causal variants), and the remaining hits PP.H1 dominance (exposure-only regional signal), consistent with the highly polygenic outcomes and the modest N of MiBioGen (per-locus posterior probabilities in Supplementary Table 5). Sample-overlap diagnostics (LDSC bivariate intercept ≤ 0.011) confirmed that overlap is not driving the signals (Supplementary Figure S7). Third, CAUSE applied to 45 taxon-outcome pairs (the five candidate clades × nine outcomes) returned zero meeting the strictest genuine-causal criterion; one pair (Negativicutes/Selenomonadales-MDD) was borderline. Despite this rejection of per-pair direct causation, CAUSE γ posterior medians were directionally concordant with the protective-versus-deleterious hypothesis in 33 of 45 pairs (sign-test binomial p = 1.23 ×10^-3^). Because several concordant γ values are close to zero, we interpret this as evidence of overall directional structure rather than as confirmation of causation (Figure 3; full CAUSE posteriors in Supplementary Table 16 and Supplementary Figure S5; an integrated cross-layer LDSC/MVMR/colocalization view in Supplementary Figure S10). Together, the discovery signals are directionally structured but not validated as direct causal effects by conservative per-locus methods.

**Figure 3.**
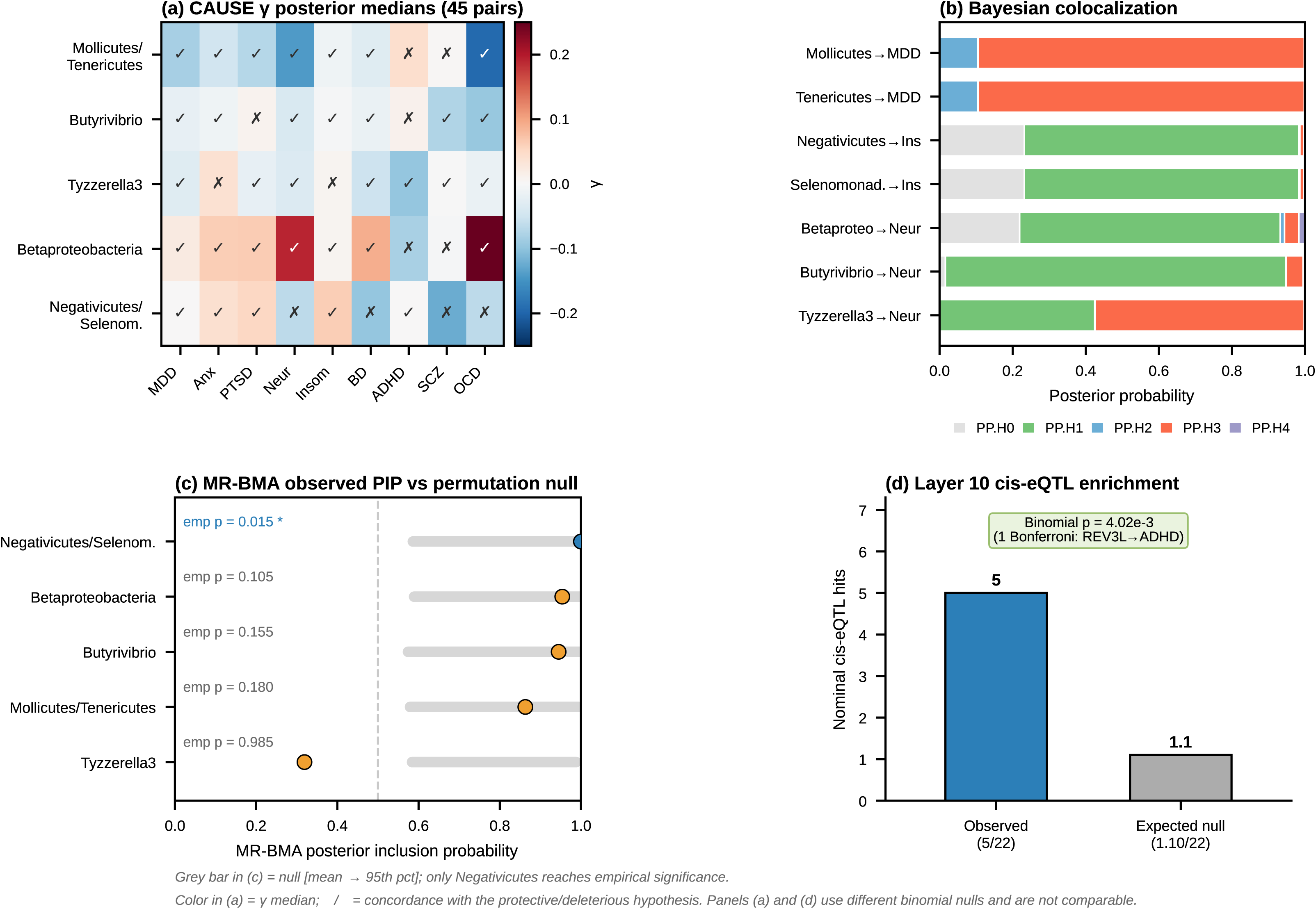
Conservative triangulation across CAUSE, Bayesian colocalization and MR-BMA. (a) CAUSE γ posterior medians for the 45 taxon-outcome pairs, annotated by directional concordance with the protective-versus-deleterious hypothesis (33/45; sign-test binomial p = 1.23 ×10^-3^). (b) Bayesian colocalization posterior probabilities (PP.H0-PP.H4) per candidate locus (PP.H4 < 0.05 for all seven; PP.H3-dominant for Mollicutes/Tenericutes-MDD and Tyzzerella3-neuroticism; PP.H1-dominant for the remaining hits). (c) MR-BMA posterior inclusion probabilities versus the permutation null (only the Negativicutes/Selenomonadales clade reaches empirical significance, p = 0.015). (d) Exploratory cis-strict cis-eQTL nominal enrichment (5/22 nominal tests; one Bonferroni- and FDR-significant signal, REV3L-ADHD). Panels (a) and (d) use different binomial null parameters and are not directly comparable.

### Selected taxa share host-genetic architecture specifically with internalizing psychopathology

The discovery concentration on internalizing outcomes motivated a latent-factor reformulation (Layer 9). The two-factor model fitted the LDSC genetic covariance matrix better than a one-factor common-psychopathology model (ΔAIC = -197), and the LDSC matrix reproduced the literature cross-disorder structure (MDD-anxiety rg = 0.91, MDD-PTSD rg = 0.93, BD-schizophrenia rg = 0.67; full genetic-correlation matrix in Supplementary Table 3 and attempted hit-taxon-outcome correlations in Supplementary Table 4). As specified in Methods, the pre-registered assignment of ADHD to the psychotic factor produced an improper solution (a Heywood case with an inflated inter-factor correlation, 0.81).

The data-consistent specification resolved this instability. With ADHD re-assigned to the internalizing/distress factor (which its genetic correlations support) and the psychotic factor defined by BD and schizophrenia, the inter-factor correlation fell to 0.48 and absolute fit improved markedly (standardized root mean residual [SRMR] 0.096 to 0.060), with a clean psychotic factor (standardized loadings BD 0.89, schizophrenia 0.75). A bifactor model, in which a general psychopathology factor and an orthogonal internalizing-specific factor are estimated jointly, retained substantial internalizing-specific loadings (0.43-0.62 for MDD, anxiety, PTSD, neuroticism and insomnia) beyond the general factor, indicating genetic variance specific to the internalizing dimension. Projected onto the internalizing-specific factor, four of the five candidate clades reached significance: Mollicutes/Tenericutes (protective; projected p = 7.7 ×10^-5^), the Negativicutes/Selenomonadales clade (deleterious; p = 7.0 ×10^-3^), Betaproteobacteria (deleterious; p = 4.8 ×10^-4^) and Butyrivibrio (protective; p = 2.6 ×10^-3^), with Tyzzerella3 nominal. None except Betaproteobacteria reached significance on the psychotic factor; Betaproteobacteria additionally showed a deleterious projection on the psychotic factor (driven by nominal effects on BD and schizophrenia), identifying it as the one broad rather than internalizing-specific taxon, consistent with the prior Betaproteobacteria-BD association reported by Ni et al. (2022). The two-factor and bifactor solutions, fit comparison and per-taxon projections are shown in Figure 4; the full model-fit comparison is in Supplementary Figure S6.

**Figure 4.**
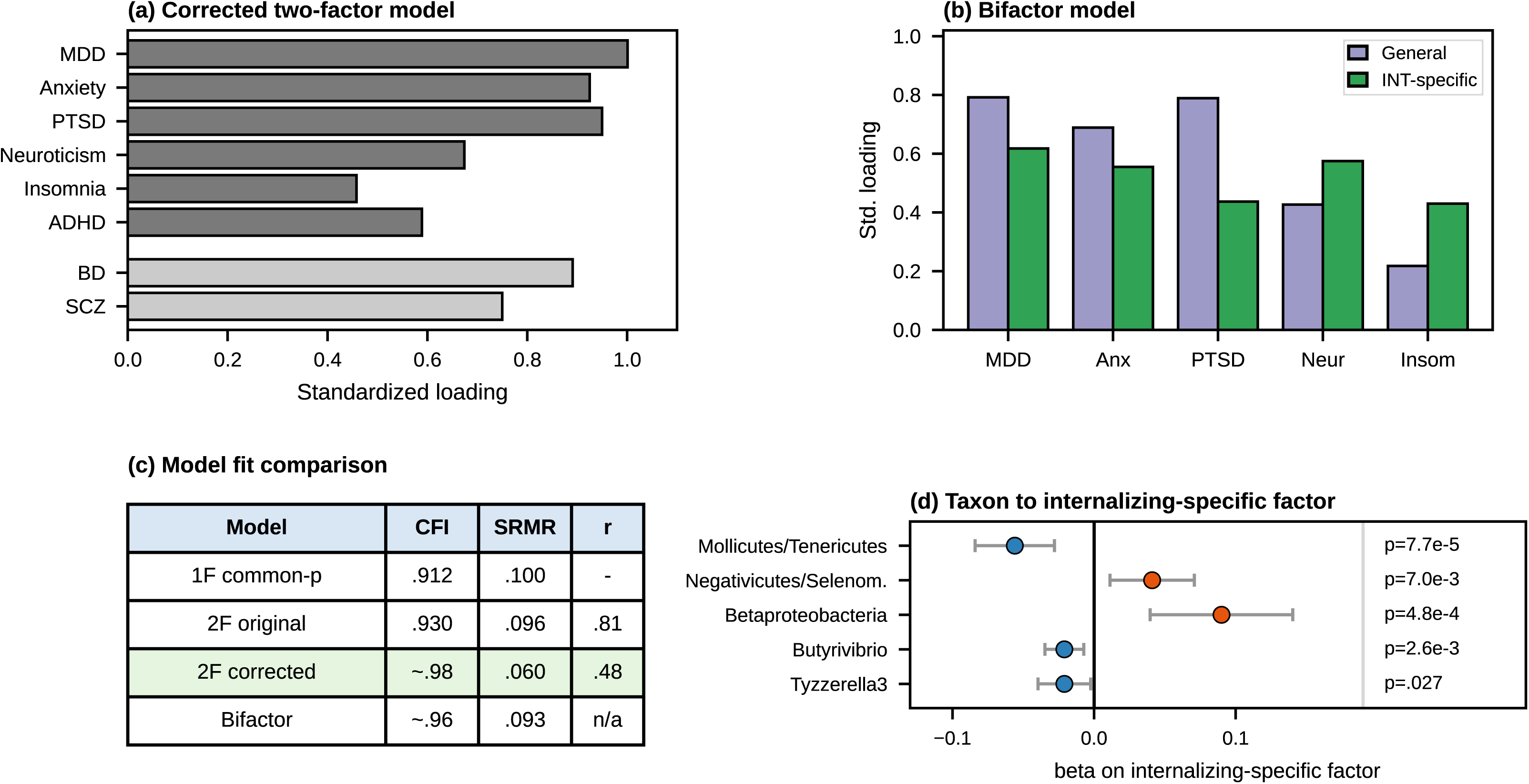
Corrected latent-factor analysis (Genomic SEM): candidate taxa share genetic architecture specifically with internalizing psychopathology. (a) Standardized loadings of the corrected two-factor model; darker grey bars are indicators of the internalizing factor (which includes ADHD on genetic grounds) and lighter grey bars the psychotic factor (BD, schizophrenia). ADHD was assigned to the internalizing/distress factor on the basis of its genetic correlations (rg with MDD 0.61 and PTSD 0.68 versus BD 0.22 and schizophrenia 0.20); the inter-factor correlation is 0.48, whereas the originally specified model placed ADHD on the psychotic factor and yielded 0.81 with a Heywood case. (b) Bifactor model with an orthogonal general psychopathology factor (purple) and an internalizing-specific factor (green); internalizing-specific genetic variance (loadings 0.43-0.62) is retained beyond the general factor. (c) Model-fit comparison across the one-factor, original two-factor, corrected two-factor and bifactor models (CFI, SRMR and inter-factor correlation r). (d) Inverse-variance-weighted projection of the candidate taxa onto the internalizing-specific factor (blue, protective β < 0; orange, deleterious β > 0; p-values shown at right); Betaproteobacteria additionally loads on the psychotic factor and is therefore broad rather than internalizing-specific. OCD was excluded from Genomic SEM for insufficient instruments. CFI and SRMR for the corrected and bifactor models are from a validated diagonally-weighted-least-squares re-fit and are being confirmed in a native Genomic SEM implementation.

Two further analyses supported and bounded this picture. MR-BMA applied to the internalizing-loading taxa, with permutation-null calibration of posterior inclusion probabilities, showed that only the Negativicutes/Selenomonadales clade achieved empirical significance (empirical p = 0.015), the other taxa being individually indistinguishable from the small-set null (Supplementary Table 17). Two-step mediation through tryptophan, kynurenine, CRP and IL-6 returned no taxon-mediator-outcome combination exceeding 5% mediation with a bootstrap CI excluding zero, indicating that the classical inflammation and tryptophan-pathway intermediates do not account for the candidate effects at current power (full estimates in Supplementary Table 2; null heatmap in Supplementary Figure S4).

### Functional annotation and exploratory cis-eQTL prioritization (hypothesis-generating)

MAGMA returned six gene-level Bonferroni hits corresponding to five named genes (ABCA7, BMI1, COMMD3, REV3L, TMEM259), but no canonical butyrate/SCFA, FFAR2/FFAR3 or HDAC pathway reached cross-taxa Bonferroni significance, so the SCFA hypothesis was not directly confirmed by formal enrichment (gene-level results in Supplementary Table 6; pathway enrichment in Supplementary Table 7). In an exploratory cis-eQTL analysis (peripheral whole-blood proxy), REV3L expression showed a Bonferroni- and FDR-significant association with ADHD (β = +0.134, p = 2.09 ×10^-3^) that lacks an obvious neurodevelopmental mechanism, and ABCA7 (a microglial lipid-transport gene) showed nominal protective associations on neuroticism and schizophrenia (instruments in Supplementary Table 8; full per-method MR results in Supplementary Table 9). We position ABCA7 and REV3L as candidate loci for brain-tissue replication and functional follow-up, not as druggable targets. These layers are hypothesis-generating and are reported in full in the Supplement.

## Discussion

This atlas integrates ten MR layers across 211 gut microbial taxa and nine psychiatric and psychopathology-related outcomes drawn from the most recent and well-powered GWAS. Three findings define the contribution. First, the directional MR signal concentrates on the internalizing dimension: every discovery hit fell on MDD, neuroticism or insomnia, and a corrected latent-factor analysis shows the candidate taxa load specifically on a latent internalizing factor and, in a bifactor model, on internalizing-specific genetic variance beyond a general psychopathology factor. Second, the most conservative criteria do not support per-locus direct causation: colocalization PP.H4 was below 0.05 at every locus, CAUSE returned zero genuinely causal pairs, and multivariable MR was over-parameterized for most hits. Third, the directional pattern is internally consistent (33/45 CAUSE γ signs concordant with the hypothesis). The most defensible interpretation is shared host-genetic architecture between selected gut microbial taxa and the genetic component of internalizing psychopathology, rather than causation from microbes to disorders at the level of individual variants (Figure 5).

**Figure 5.**
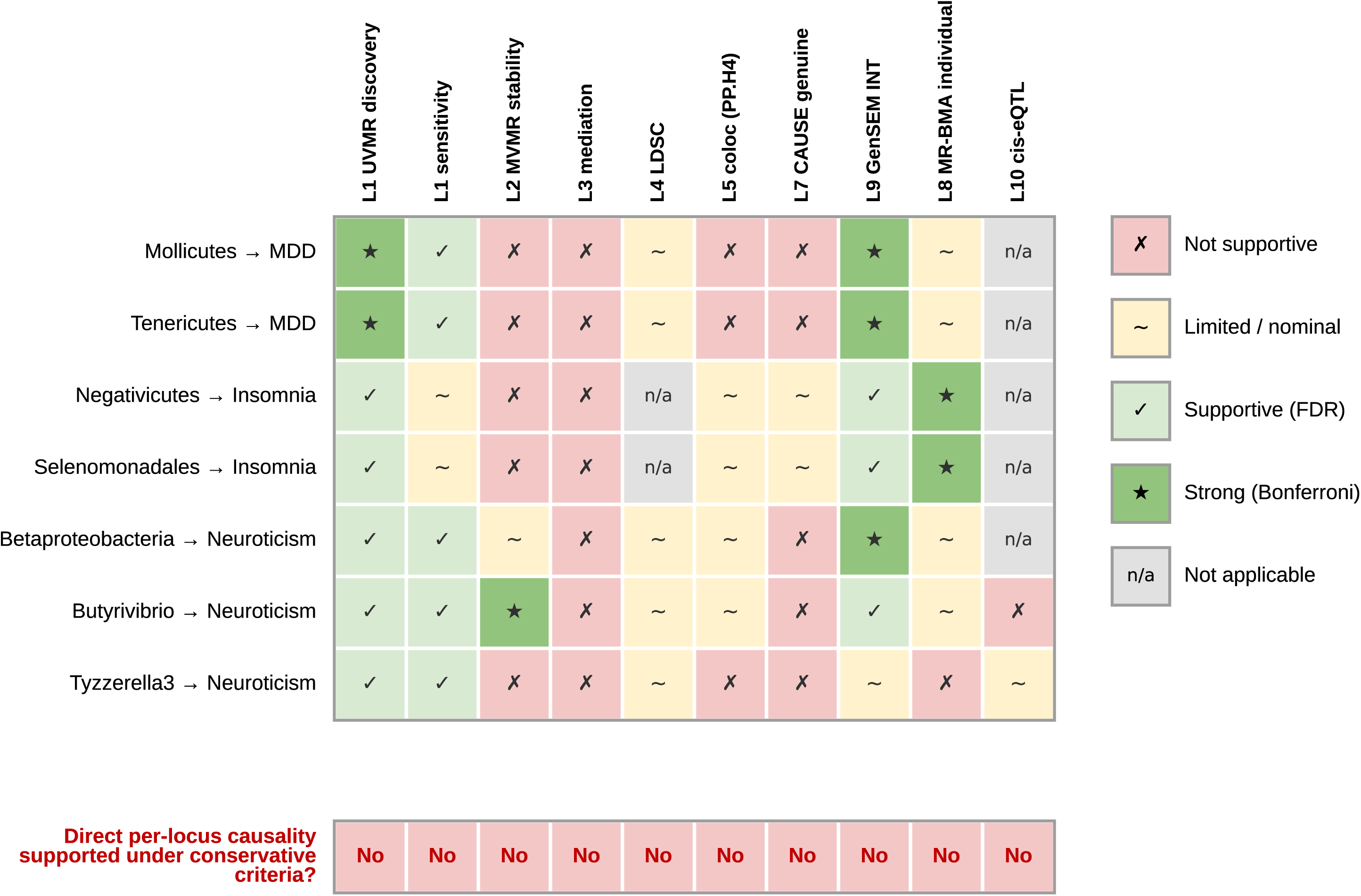
Evidence triangulation matrix per candidate hit. Per-hit evidence symbol across the ten analytical layers (filled star, Bonferroni-strong; check, FDR-supportive; tilde, limited/nominal; cross, not supportive; n/a, not applicable). The bottom row indicates that none of the seven candidate hits passes the strictest per-locus criteria (CAUSE genuine-causal with γ credible interval excluding zero; colocalization PP.H4 > 0.8). Layer 10 yielded one Bonferroni- and FDR-significant cis-eQTL signal (REV3L-ADHD), but ADHD is not among the candidate hits’ outcomes. The converging directional signal across layers supports shared host-genetic architecture with the internalizing dimension rather than direct per-locus causation.

### Comparison with previous MR studies of gut microbiome and psychiatry

The closest comparators applied broad MR scans with MiBioGen. Wu et al. (2025) examined six disorders (ADHD, anxiety, BD, anorexia, schizophrenia, autism) but did not include MDD, OCD, PTSD, insomnia or neuroticism, and reported mediation through M-CSF, GDNF and CD40 that is orthogonal to our pre-specified inflammation/tryptophan mediators; their anxiety GWAS (∼2,000 cases) is two orders of magnitude smaller than the release used here (122,341 cases), which helps explain why anxiety did not reach FDR in our better-powered analysis. Ni et al. (2022) applied univariable MR to 190 taxa across 15 outcomes and identified class Betaproteobacteria as deleterious for BD; this is the one signal that converges with our atlas, where Betaproteobacteria is deleterious on neuroticism and is the single taxon loading on both the internalizing and psychotic factors. Xiao et al. (2023) used METAL/MTAG-augmented microbiome GWAS and reported species-level Bacteroides hits not resolvable by the genus-level MiBioGen V4 instrument set, explaining the absence of overlap with our genus/class/order-level hits. Ye et al. (2025) used brain-structural endophenotypes as mediators, a complementary axis to our circulating mediators, and the two studies converge on a microbiota-brain-axis model while identifying non-overlapping specific taxa. Thus the literature is internally heterogeneous at the level of individual taxa (no taxon replicates across all four scans), but homogeneous at a higher level: butyrate-related/Firmicutes signals recur as protective and Proteobacteria-related signals as deleterious, and Betaproteobacteria is the most reproducible single candidate. None of these four applied CAUSE, permutation-null MR-BMA, or a latent-factor decomposition to taxon-disorder pairs; the comparative contribution of our work is therefore the conservative cascade and the dimensional reframing more than the discovery of new taxa.

### Mechanism, methodological contributions and the value of a corrected null

The canonical SCFA hypothesis is most relevant to the genus-level protective signals (Butyrivibrio, Tyzzerella3), but no SCFA, FFAR2/FFAR3 or HDAC pathway reached cross-taxa Bonferroni significance, so it cannot be cited as confirmed; convergent nominal enrichment instead implicates epigenetic, immune, neurogenesis and microglial processes that require direct experimental adjudication. Two methodological elements may be useful independently of the biology: the diagnosis and correction of standard-error inflation in multivariable MR with weak microbial instruments; and permutation-null calibration of MR-BMA posterior inclusion probabilities. We also illustrate the importance of grounding latent-factor specifications in the genetic correlation structure rather than clinical nosology: a single mis-assignment (ADHD) inflated an inter-factor correlation and produced an improper solution, whereas a data-consistent re-specification and a bifactor model yielded a stable, interpretable internalizing-specific signal. Finally, by applying conservative per-locus tests uniformly, we provide a stringent test of the direct-causation hypothesis and find it negative, while the dimensional-architecture finding remains positive and interpretable.

### The atlas as a public resource

Although our narrative is focused on the internalizing dimension, the full atlas (all 211 × 9 univariable tests and all ten layers) is reported in the Supplement and deposited on OSF, so that the complete analytic landscape, including null layers, is available to the field and protected against selective reporting. The seven candidate taxa and the ABCA7 and REV3L loci are priorities for follow-up in larger microbiome GWAS, brain-tissue cis-eQTL analyses and direct experimental validation.

## Limitations

Several limitations apply. Horizontal pleiotropy cannot be fully excluded, although MR-Egger, weighted median, MR-PRESSO and CAUSE were applied. Weak-instrument bias remains a concern given the p < 1 ×10^-5^ threshold required by MiBioGen sample size; mean F > 19 and MR-RAPS mitigate but do not eliminate it, and winner’s curse may inflate effect sizes. The sign test summarising directional concordance across the 45 taxon-outcome pairs assumes independence, which does not hold because five clades were tested against nine genetically correlated outcomes; its p-value should therefore be read as descriptive rather than as a formal test. Analyses were restricted to European- ancestry data and may not generalize. The MiBioGen V4 instrument set resolves taxa to genus but not species/strain. Bayesian colocalization assumed a single causal variant; multi-causal fine-mapping (coloc-SuSiE) would require LD matrices not available for MiBioGen. The Genomic SEM analysis is exploratory and was re-fitted after a declared correction; it contains a residual near-Heywood loading for MDD (reflecting near-saturation of the MDD-internalizing communality) and the candidate taxa were identified from internalizing-domain associations, so the latent-factor projection supports but does not independently prove preferential internalizing alignment. The bifactor model should be confirmed in a native Genomic SEM implementation with full standard errors. Mediation depended on mediator instruments with smaller GWAS than the outcomes. Layer 10 is an exploratory peripheral-blood proxy requiring brain-tissue replication. Finally, the outcome panel does not cover the full DSM-5 nosology, and two phenotypes (neuroticism, insomnia) are trait/symptom rather than diagnostic categories.

## Data Availability

All data analysed are summary-level statistics from previously published genome-wide association studies, listed with full provenance, accessions and download URLs in Supplementary Table 1. Harmonized datasets, analysis code (including the corrected Genomic SEM re-analysis) and intermediate outputs are deposited on the Open Science Framework (DOI 10.17605/OSF.IO/FVGQ2). Analysis code is also available at https://github.com/pedrovelezpardo/mr-microbiome-psychiatric-spectrum.

https://doi.org/10.17605/OSF.IO/FVGQ2

https://github.com/pedrovelezpardo/mr-microbiome-psychiatric-spectrum

https://mibiogen.gcc.rug.nl/

https://pgc.unc.edu/for-researchers/download-results/

https://gwas.mrcieu.ac.uk/

## Data and code availability

Genetic summary statistics are publicly available or consortium-accessible from the repositories listed in Supplementary Table 1. Harmonized datasets, analysis code (including the corrected Genomic SEM re-analysis) and intermediate outputs are deposited on the Open Science Framework (DOI 10.17605/OSF.IO/FVGQ2).

## Funding

This research did not receive any specific grant from funding agencies in the public, commercial, or not-for-profit sectors. The work was conducted with self-funded doctoral resources; the authors’ institutions had no role in study design, in the collection, analysis and interpretation of data, in the writing of the report, or in the decision to submit the article for publication.

## CRediT authorship contribution statement

Pedro Velez-Pardo: Conceptualization, Methodology, Software, Formal analysis, Data curation, Visualization, Investigation, Writing – original draft, Writing – review & editing, Project administration. Roger Jose Solano: Conceptualization, Methodology, Investigation, Resources, Supervision, Writing – review & editing. Adriana Maria Quinchia-Figueroa: Conceptualization, Resources, Supervision, Writing – review & editing. Ricardo Montoya Monsalve: Data curation, Investigation, Writing – review & editing. Nadia Semenova Moratto-Vasquez: Conceptualization, Methodology, Resources, Supervision, Writing – review & editing. All authors read and approved the final manuscript.

## Declaration of competing interest

The authors declare that they have no known competing financial interests or personal relationships that could have appeared to influence the work reported in this paper.

## Ethics approval

This study is a secondary analysis of publicly available or consortium-accessible, de-identified, aggregate genome-wide association summary statistics (sources in Supplementary Table 1). Ethics approval and informed consent were obtained by the original studies that generated each dataset; no new individual-level data were collected and no additional ethics committee approval was required for this work.

## Acknowledgements

We thank the MiBioGen consortium and the Psychiatric Genomics Consortium working groups for making their summary statistics publicly available; the contributing investigators of the Adams 2025 (MDD), Strom 2026 (anxiety), Strom 2025 (OCD), Mullins 2021 (BD), Trubetskoy 2022 (schizophrenia), Nievergelt 2024 (PTSD) and Demontis 2023 (ADHD) studies; the MRC-IEU OpenGWAS infrastructure for hosting harmonized UK Biobank data; and the eQTLGen consortium and DGIdb maintainers for cis-eQTL and pharmacological annotation resources.

## Declaration of generative AI and AI-assisted technologies in the manuscript preparation process

During the preparation of this work the authors used Claude (Anthropic) in order to draft and language-edit the manuscript; use of the same assistant to reproduce and cross-check the Genomic SEM re-analysis and to generate plotting code for the display items is described in the Methods. After using this tool, the authors reviewed and edited the content as needed and take full responsibility for the content of the published article.

## Notes

### Competing Interest Statement

The authors have declared no competing interest.

### Clinical Protocols

https://doi.org/10.17605/OSF.IO/FVGQ2

### Author Declarations

This study used only publicly available, de-identified, aggregate GWAS summary statistics. No individual-level data were accessed. Ethics approval and informed consent were obtained by the original studies that generated each dataset; therefore, no additional ethics committee approval was required for this secondary analysis of openly available human summary data.

### Summary of Updates

This version updates the manuscript as follows. Abstract: the contrast for the FDR-significant findings now reads "rather than on bipolar disorder or schizophrenia". ADHD was removed from that contrast because it was inconsistent with the corrected Genomic SEM reported in this manuscript, in which ADHD is assigned to the internalizing factor on genetic grounds. Non-standard abbreviations are now defined at first mention: the colocalization posterior probability of a shared causal variant (PP.H4) and the summary-data causal test (CAUSE). No numerical result changed. Methods: the latent-factor section no longer states that a three-factor structure was compared; the models actually fitted and reported are one-factor, two-factor and bifactor. The reassignment of ADHD to the internalizing factor is now reported as a data-driven deviation from the pre-registered factor structure, documented as a declared amendment on OSF, rather than as a post-review correction; the corrected model is presented as the primary model. A subsection describing the use of an AI assistant in the analysis has been added, and a declaration of generative AI use in manuscript preparation has been added before the references. Ethics: a dedicated ethics approval statement has been added, stating that the study is a secondary analysis of publicly available or consortium-accessible, de-identified, aggregate GWAS summary statistics, that ethics approval and informed consent were obtained by the original studies, and that no additional ethics committee approval was required. Results: the CAUSE analysis is now explicitly described as covering 45 taxon-outcome pairs, corresponding to the five candidate clades tested against nine outcomes. Limitations: a caveat has been added noting that the sign test summarising directional concordance across the 45 taxon-outcome pairs assumes independence, which does not hold because five clades were tested against nine genetically correlated outcomes; its p-value should be read as descriptive rather than as a formal test. Author affiliations updated: the corresponding author is now affiliated only with Universidad EIA, where the work was carried out. Co-author affiliations are unchanged. Formatting: references and in-text citations have been converted to author-year style, thousands separators are now commas, and Table 1 is embedded as editable text. No data, estimates or conclusions were changed.

