## Supplementary material for "Shared host-genetic architecture between gut microbiota and internalizing psychopathology" (from another version of this preprint: version 1, 2026-06-02)

### Supplementary Information — guide, titles and file summaries

*Gut microbial taxa share host-genetic architecture with the internalizing dimension of psychopathology: a pre-registered Mendelian randomization atlas.*

Supplementary information is available at Molecular Psychiatry’s website. Each item below is referred to at least once in the main text. Tables are provided as .csv and .xlsx; figures as vector .pdf/.eps with .png/.tiff copies. Numbering is continuous.

#### Supplementary Tables

##### Supplementary Table 1. *Genome-wide association study data sources.*

Provenance of every dataset used — the nine psychiatric/psychopathology outcomes, four mediators and the MiBioGen exposure: consortium/first author, year, phenotype definition, total/case/control sample sizes, ancestry, UK Biobank and 23andMe inclusion, genome build, accession identifiers, download URLs and dates.

*File format: .csv and .xlsx*

##### Supplementary Table 2. *Two-step mediation results.*

Complete two-step mediation for the seven candidate hits through four pre-specified mediators (tryptophan, kynurenine, CRP, IL-6): 28 taxon–mediator–outcome combinations with indirect-effect estimates, 10,000-bootstrap confidence intervals and proportion mediated. No combination exceeded 5% with an interval excluding zero.

*File format: .csv and .xlsx*

##### Supplementary Table 3. *LDSC genetic-correlation matrix among outcomes.*

Linkage-disequilibrium-score-regression genetic correlations (with standard errors) among the nine outcomes, reproducing the established cross-disorder structure and providing the genetic covariance matrix for the latent-factor (Genomic SEM) analysis.

*File format: .csv and .xlsx*

##### Supplementary Table 4. *Attempted LDSC genetic correlations: hit taxa versus outcomes.*

The 63 attempted genetic correlations between candidate hit taxa and the nine outcomes. Most were non-estimable because MiBioGen taxon heritability is very low, as expected for 16S-based microbial traits.

*File format: .csv and .xlsx*

##### Supplementary Table 5. *Bayesian colocalization posterior probabilities.*

Per-candidate-locus coloc.abf results: top SNP, chromosome, position, window and posterior probabilities PP.H0–PP.H4 with interpretation. PP.H4 was below 0.05 for all seven hits, indicating no shared single causal variant.

*File format: .csv and .xlsx*

**Supplementary Table 6.** ***MAGMA gene-level results.***

Gene-level MAGMA statistics for the candidate taxa: gene symbol, chromosome, position, SNP count, Z-statistic, p-value, taxon and Bonferroni-significance flag. Six gene-level Bonferroni hits correspond to five named genes.

*File format: .csv and .xlsx*

**Supplementary Table 7.** ***MAGMA gene-set/pathway enrichment (nominal).***

Nominally significant ($p<0.05$) MAGMA gene-set enrichments across taxa: gene-set name, type, gene count, effect size, standard error, p-value, source database and Bonferroni flag. No pathway reached cross-taxa Bonferroni significance.

*File format: .csv and .xlsx*

**Supplementary Table 8.** ***Cis-strict cis-eQTL instruments per drug-target gene.***

Whole-blood (eQTLGen) cis-eQTL instruments used in Layer 10, per target gene: SNP, chromosome, position, alleles, effect-allele frequency, effect size, standard error, p-value, sample size and target gene, after cis-strict filtering.

*File format: .csv and .xlsx*

**Supplementary Table 9.** ***Cis-eQTL Mendelian randomization, full results.***

All Layer-10 gene-to-outcome MR results: 22 IVW tests across three genes (ABCA7, REV3L, TMEM259) and nine outcomes, each with IVW, MR-Egger and weighted-median estimates (66 rows): effect, standard error, p-value and SNP count.

*File format: .csv and .xlsx*

**Supplementary Table 10.** ***Univariable MR sensitivity estimates for the seven hits.***

IVW, MR-Egger and weighted-median effect sizes, SNP counts and p-values for the seven candidate taxon–outcome hits, documenting direction concordance across estimators.

*File format: .csv and .xlsx*

**Supplementary Table 11.** ***Cochran's Q heterogeneity for the seven hits.***

Cochran's Q heterogeneity statistics (IVW) for the seven candidate hits: SNP count, degrees of freedom and p-value. All non-significant.

*File format: .csv and .xlsx*

**Supplementary Table 12.** ***MR-Egger intercept (directional-pleiotropy) test.***

MR-Egger intercept directional-pleiotropy test for the seven candidate hits: SNP count and intercept p-value. All intercepts non-significant.

*File format: .csv and .xlsx*

**Supplementary Table 13.** ***Per-taxon instrument strength (F-statistics).***

Instrument strength for all 211 MiBioGen taxa: SNPs before and after LD-clumping and mean F-statistic per taxon (range 20.3–25.9, median 21.6), supporting adequate but threshold-limited instrument strength.

*File format: .csv and .xlsx*

**Supplementary Table 14.** ***Layer-2 multivariable (cross-disorder) MR, taxon-only specification.***

Multivariable MR estimates (taxon-only-instrument PRIMARY specification) for each candidate hit across the eight-outcome panel: effect size, standard error, t-value and p-value per taxon–outcome.

*File format: .csv and .xlsx*

**Supplementary Table 15.** ***Layer-2 multivariable MR variance-inflation diagnostics.***

Variance-inflation factors for the PRIMARY and sensitivity multivariable-MR specifications, documenting structural over-parameterization (taxon VIF = infinite or >700) for five of seven hits and the resulting limited interpretability.

*File format: .csv and .xlsx*

**Supplementary Table 16.** ***CAUSE posteriors for 45 taxon–outcome pairs.***

Full CAUSE output for the 45 pairs (five unique hit taxa × nine outcomes): gamma, eta and q posterior medians with credible intervals, causal-versus-sharing comparison (z, p, ELPD) and conclusion category. Zero pairs were genuinely causal.

*File format: .csv and .xlsx*

**Supplementary Table 17.** ***MR-BMA posterior inclusion probabilities with permutation null.***

Marginal posterior inclusion probabilities from MR-BMA for the internalizing-loading taxa, with permutation-null mean, 95th percentile and empirical p-values. Only the Negativicutes/Selenomonadales clade reached empirical significance.

*File format: .csv and .xlsx*

#### Supplementary Figures

**Supplementary Figure S1.** ***Instrument selection and harmonization flow.***

Flow diagram of the instrument pipeline for the MiBioGen exposures: SNP selection at $p<1\times10^{-5}$, LD-clumping, palindromic-SNP exclusion and re-harmonization with each outcome GWAS.

*File format: .pdf/.eps (vector) + .png/.tiff*

**Supplementary Figure S2.** ***Univariable MR p-value landscape (211 × 9).***

Heatmap of inverse-variance-weighted MR p-values across all 211 taxa and nine outcomes (1,898 valid tests), showing the full discovery landscape from which the seven FDR hits emerge.

*File format: .pdf/.eps (vector) + .png/.tiff*

**Supplementary Figure S3.** ***Distribution of per-taxon instrument F-statistics.***

Histogram of mean F-statistics across the 211 MiBioGen taxa (median 21.6, range 20.3–25.9), illustrating the narrow strength range imposed by the $p<1\times10^{-5}$ instrument threshold.

*File format: .pdf/.eps (vector) + .png/.tiff*

**Supplementary Figure S4.** ***Mediation: null proportion-mediated heatmap.***

Heatmap of proportion-mediated estimates for the seven hits across four mediators; all combinations null (|proportion|<5% with bootstrap confidence intervals including zero).

*File format: .pdf/.eps (vector) + .png/.tiff*

**Supplementary Figure S5.** ***CAUSE posterior distributions (45 pairs).***

Posterior distributions of the causal parameter for all 45 CAUSE taxon–outcome pairs, showing that none meets the genuine-causal criterion (credible interval excluding zero with a favoured causal model).

*File format: .pdf/.eps (vector) + .png/.tiff*

**Supplementary Figure S6.** ***Genomic SEM model-fit comparison.***

Comparison of one-factor, original and corrected two-factor, and bifactor Genomic SEM solutions (fit indices and loadings), supporting the corrected internalizing-specific specification.

*File format: .pdf/.eps (vector) + .png/.tiff*

**Supplementary Figure S7.** ***Sample-overlap diagnostic (LDSC bivariate intercept).***

LDSC bivariate (gcov) intercepts for the estimable hit pairs, all ≤0.011, confirming negligible sample overlap between exposure and outcome datasets.

*File format: .pdf/.eps (vector) + .png/.tiff*

**Supplementary Figure S8.** ***Volcano plot of univariable MR.***

Volcano plot of the 1,898 univariable MR tests (effect size versus −log10 p-value), highlighting the seven FDR-significant hits, all on internalizing outcomes.

*File format: .pdf/.eps (vector) + .png/.tiff*

**Supplementary Figure S9.** ***Focused forest plot of the seven candidate hits.***

Forest plot of the seven candidate taxon–outcome hits across IVW, MR-Egger and weighted-median estimators, showing direction concordance and effect-size stability.

*File format: .pdf/.eps (vector) + .png/.tiff*

**Supplementary Figure S10.** ***Integrated cross-layer view (LDSC, MVMR, colocalization).***

Four-panel cross-validation: (a) LDSC genetic-correlation heatmap among outcomes; (b) hit-taxon–outcome genetic correlations with 95% confidence intervals; (c) Layer-2 multivariable-MR estimates for the seven hit pairs; (d) colocalization posterior probabilities per hit pair.

*File format: .pdf/.eps (vector) + .png/.tiff*

**Figure S1 | Instrument selection flow (CONSORT-like) for the seven Layer 1 candidate hits**

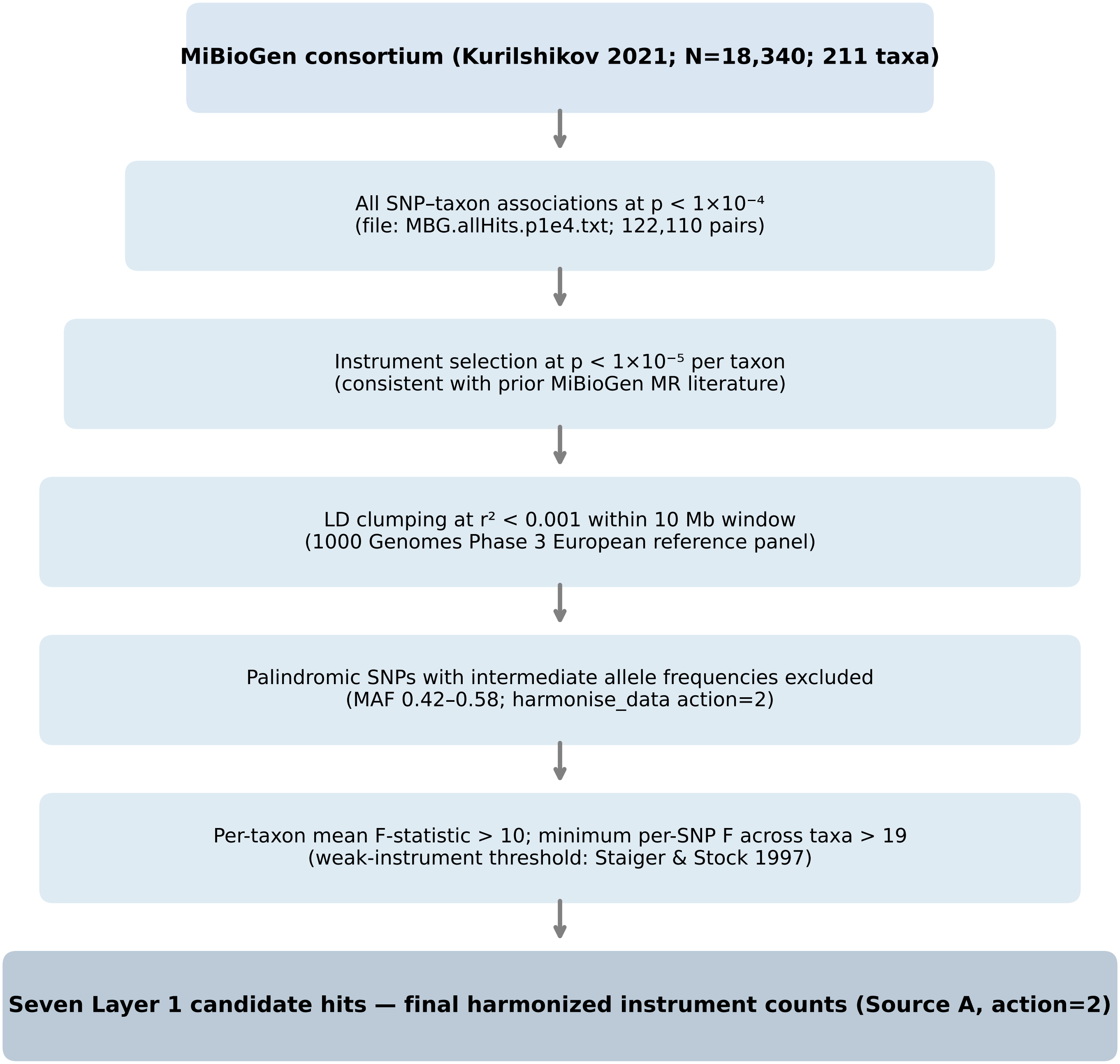


| Taxon | Outcome | n SNPs | Mean F |
|---|---|---|---|
| c. Mollicutes / p. Tenericutes | MDD | 11 | 21.6 |
| c. Negativicutes / o. Selenomonadales | Insomnia | 12 | 20.3 |
| c. Betaproteobacteria | Neuroticism | 12 | 22.8 |
| g. Butyrivibrio | Neuroticism | 15 | 22.4 |
| g. Tyzzerella3 | Neuroticism | 13 | 25.9 |

*Note: hits in rows 1 and 2 correspond to single biological clades represented redundantly at two taxonomic ranks. After deduplication, the seven hits correspond to five effective biological clades.*

**Supplementary Fig S2 | Full 211 × 9 UVMR p-value heatmap (n=211 taxa)**

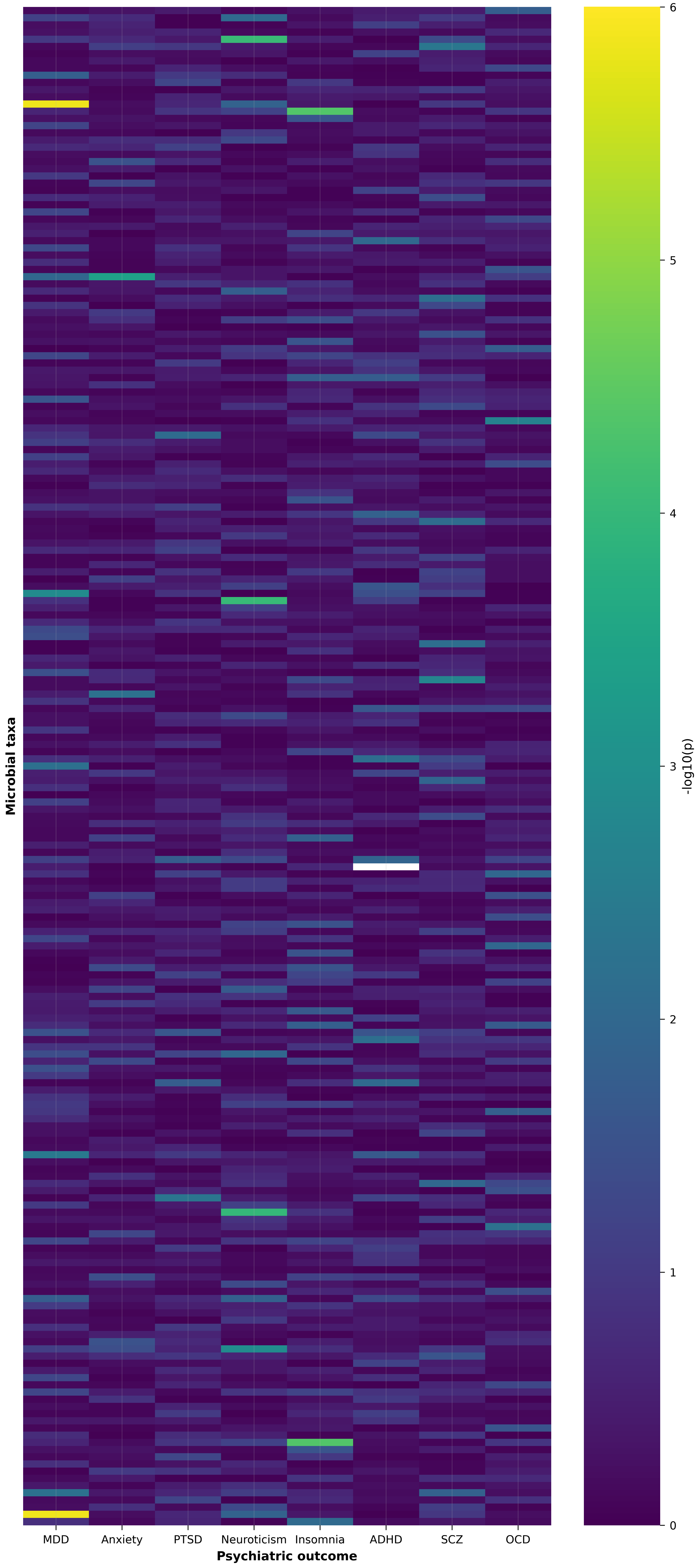

**Figure S3 | Instrument strength across 211 MiBioGen microbial taxa**

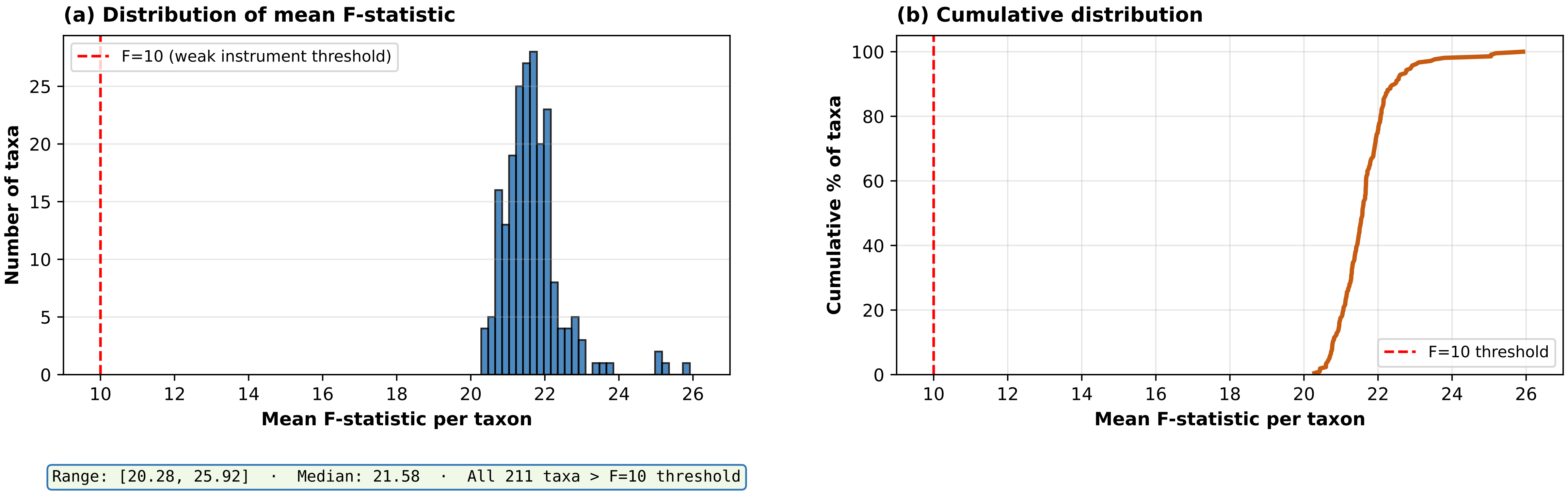


*Per-taxon mean F-statistic computed on retained instrumental SNPs after LD-clumping at $r^2 < 0.001$ within 10 Mb (1000 Genomes Phase 3 European reference). All 211 taxa exceed the F=10 weak-instrument threshold (Staiger & Stock 1997).*

**Figure S4 | Layer 3 two-step mediation — proportion mediated (%) for the 7 candidate hits × 4 mediators**

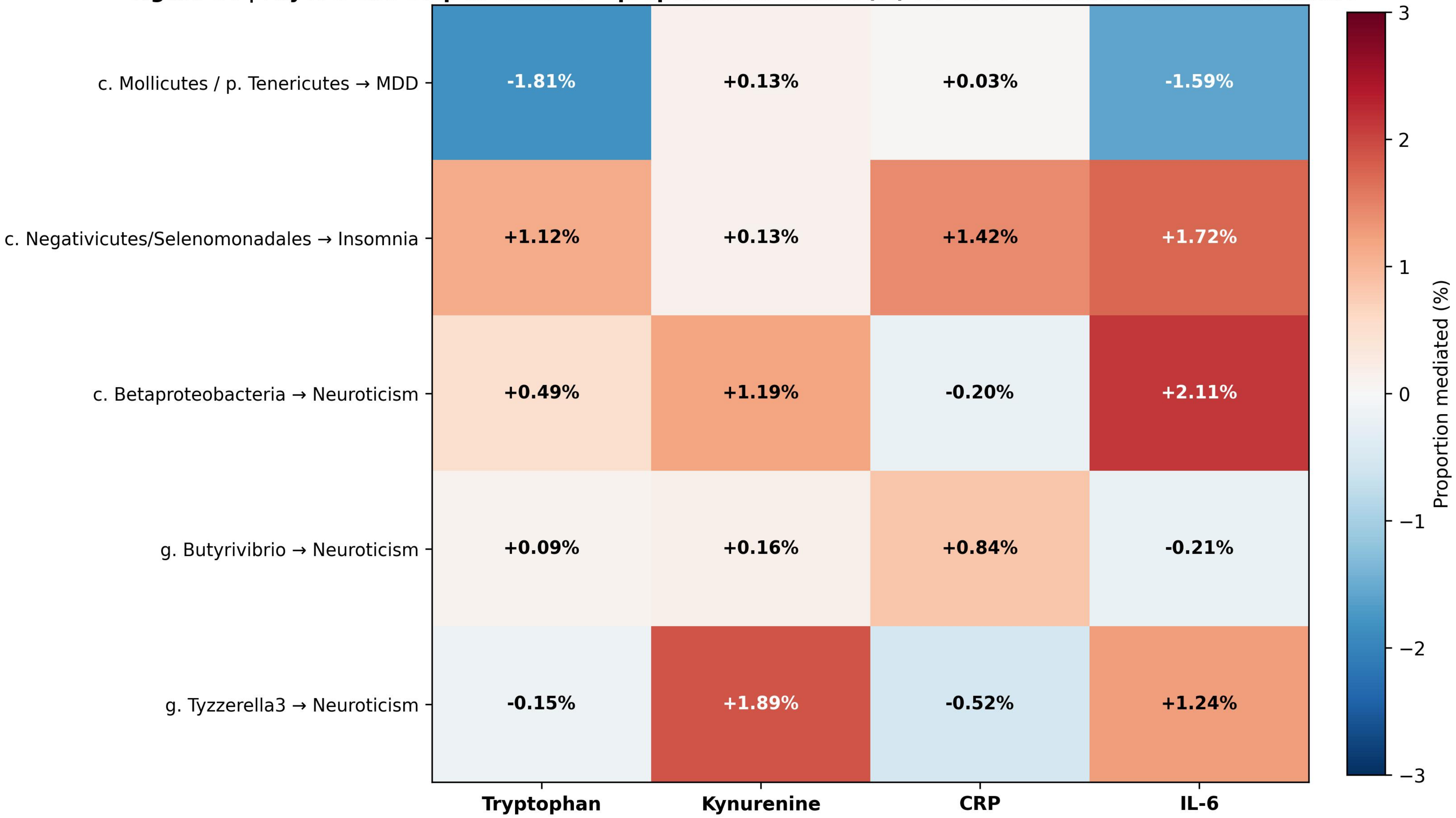


*Values from Supplementary Table 2 (Layer3_mediation_results.csv). All 28 taxon–mediator–outcome combinations: indirect_p > 0.37, |proportion mediated| < 2.1%, bootstrap 95% CI crossing zero. Mediation through CRP, IL-6, tryptophan or kynurenine is null at available statistical power.*

**Figure S5 | Layer 7 CAUSE — full posterior medians for the 45 evaluated taxon–outcome pairs**

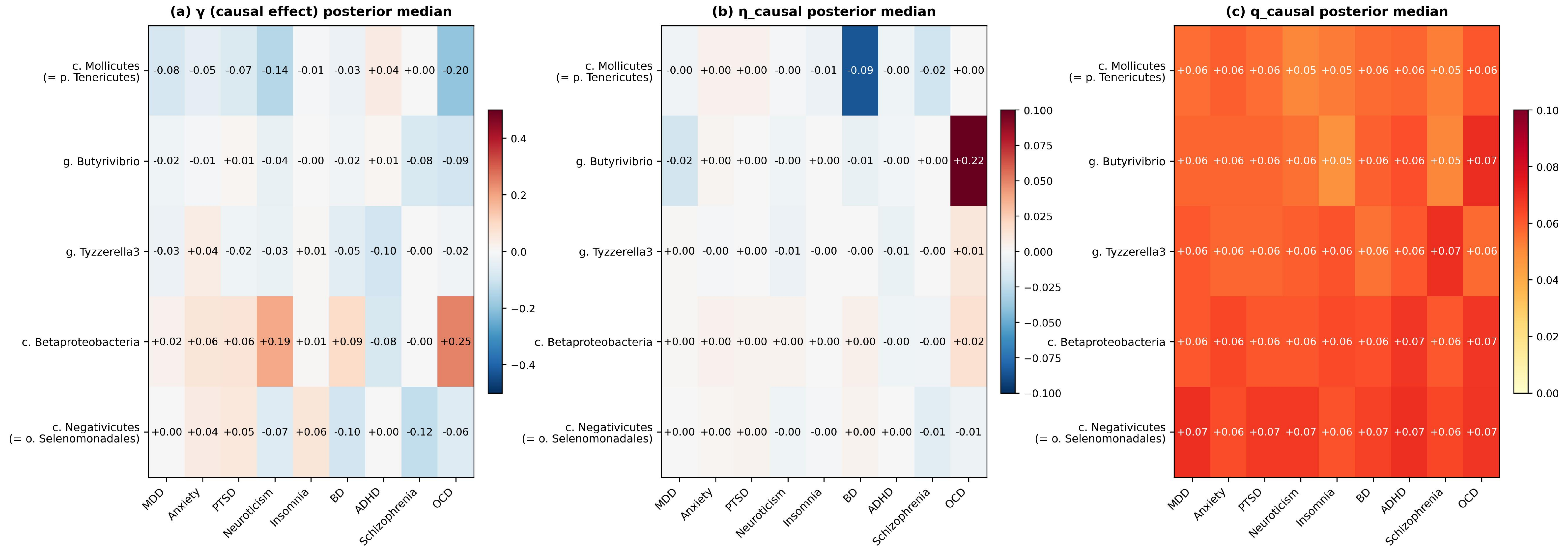


*0/45 pairs meet the CAUSAL_GENUINO criterion; 1/45 BORDERLINE (Negativicutes/Selenomonadales → MDD); 44/45 NO_EFFECT.*
*33/45 pairs show γ-sign concordant with the protective-vs-deleterious taxonomic hypothesis (binomial sign-test $p = 1.23\times10^{-3}$, one-sided exact under $p_{null}$=0.5).*

**Figure S6 | Genomic SEM model comparison: one-factor, two-factor (original / corrected) and bifactor**

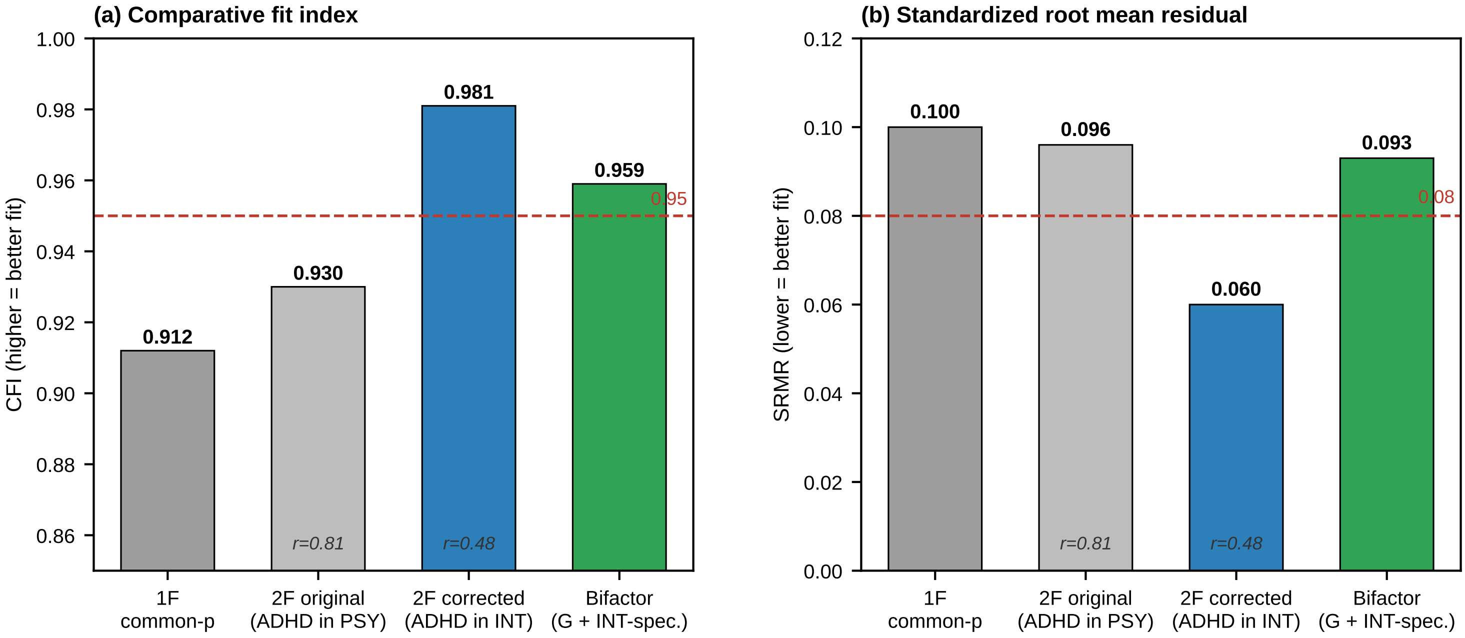


DWLS estimation on the LDSC genetic covariance matrix (OCD excluded). The corrected two-factor model (ADHD re-assigned to the internalizing factor) and the bifactor model are the finalised specifications; r = inter-factor correlation (0.81 in the mis-specified original, 0.48 corrected). CFI/SRMR for corrected and bifactor are from a validated DWLS re-fit (confirm in native GenomicSEM).

Figure S7 | Sample overlap between MiBioGen and the seven candidate hit outcomes (LDSC gcov_int)

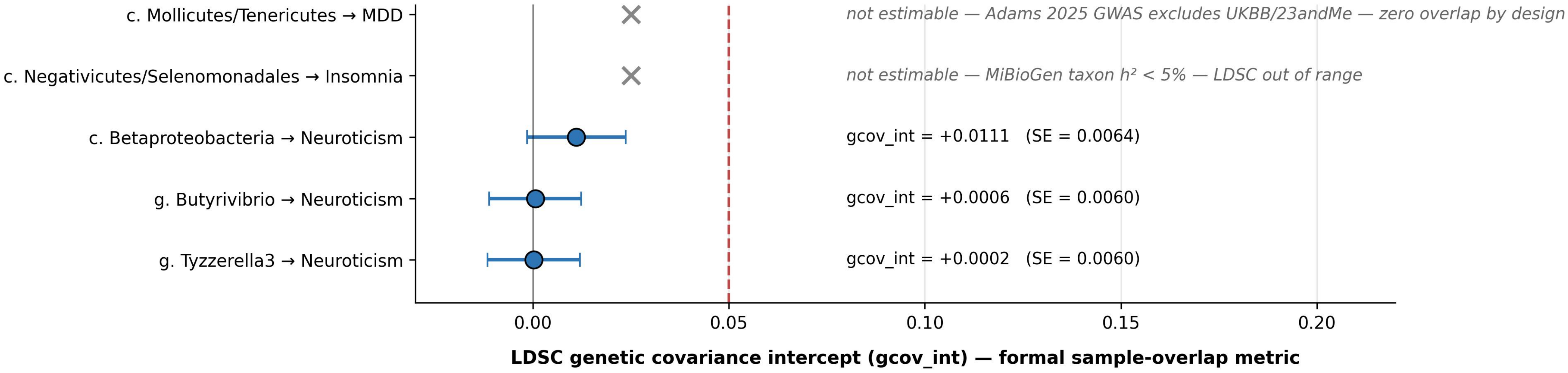


*All three estimable gcov_int values are ≤ 0.011, well below the 0.05 concern threshold (Bulik-Sullivan et al. 2015).*
*For the Mollicutes/Tenericutes → MDD hit, the Adams 2025 GWAS explicitly excludes UKBB and 23andMe, so zero sample overlap is expected by design.*
*For the Negativicutes/Selenomonadales → Insomnia hit, LDSC could not be estimated due to low MiBioGen heritability ($h^2 < 5\%$) for that taxon.*
*Formal sample-overlap corrections (MRlap) were therefore not required for Layer 1.*

**Figure S8 | UVMR discovery landscape across 1,898 taxon-outcome tests**

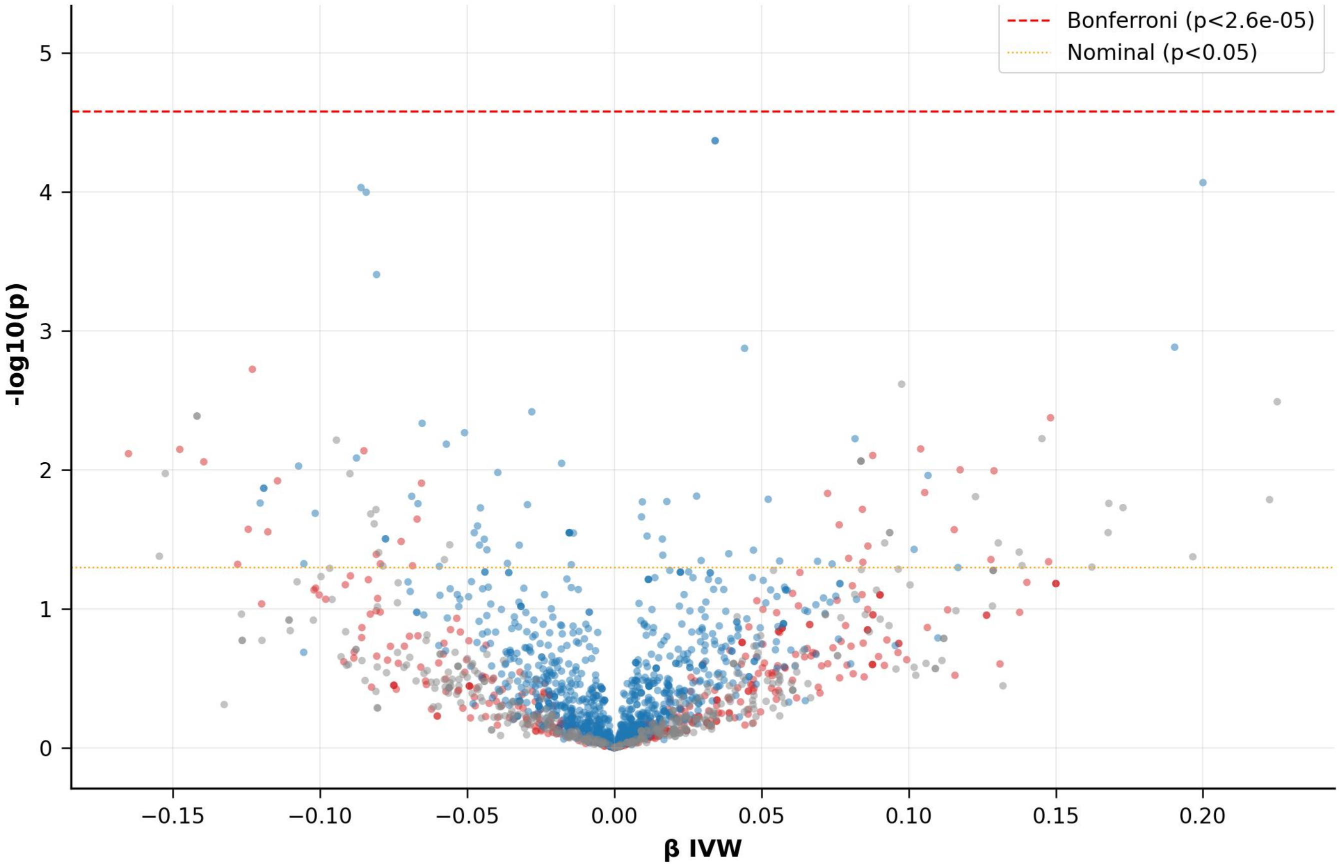

**Figure S9 | Focused forest plot — seven Layer 1 candidate hits across 3 MR methods (Source A, action=2 harmonization)**

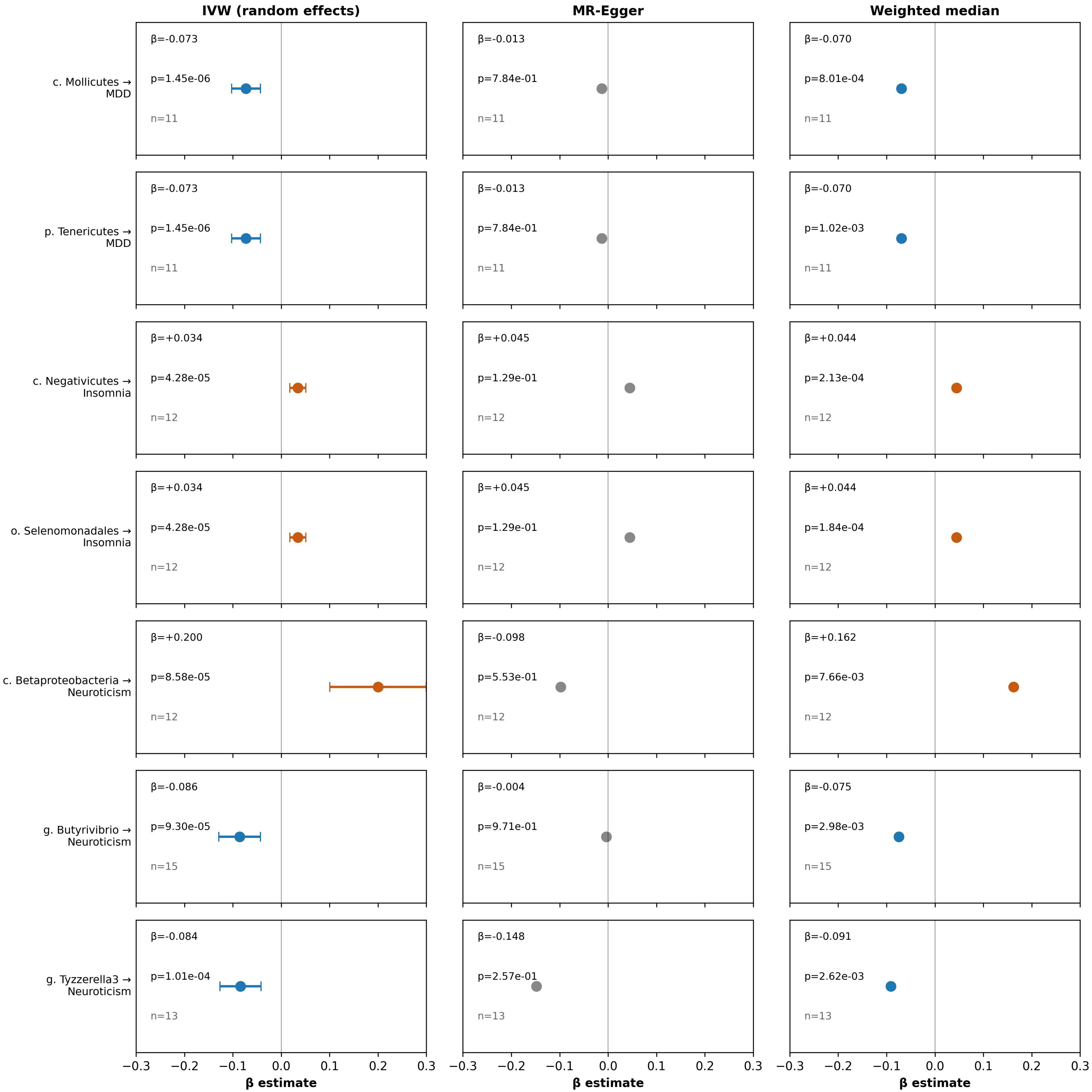


*All estimates from Layer1_UVMR_results.csv (action=2 harmonization). Cochran Q p-values in ST10; MR-Egger intercept p in ST11.*
*Weighted mode not reported here (not in primary Source A); future work with supplementary manual_harmonise can add.*

**Figure S10 | Cross-validation — LDSC + MVMR + Colocalization**

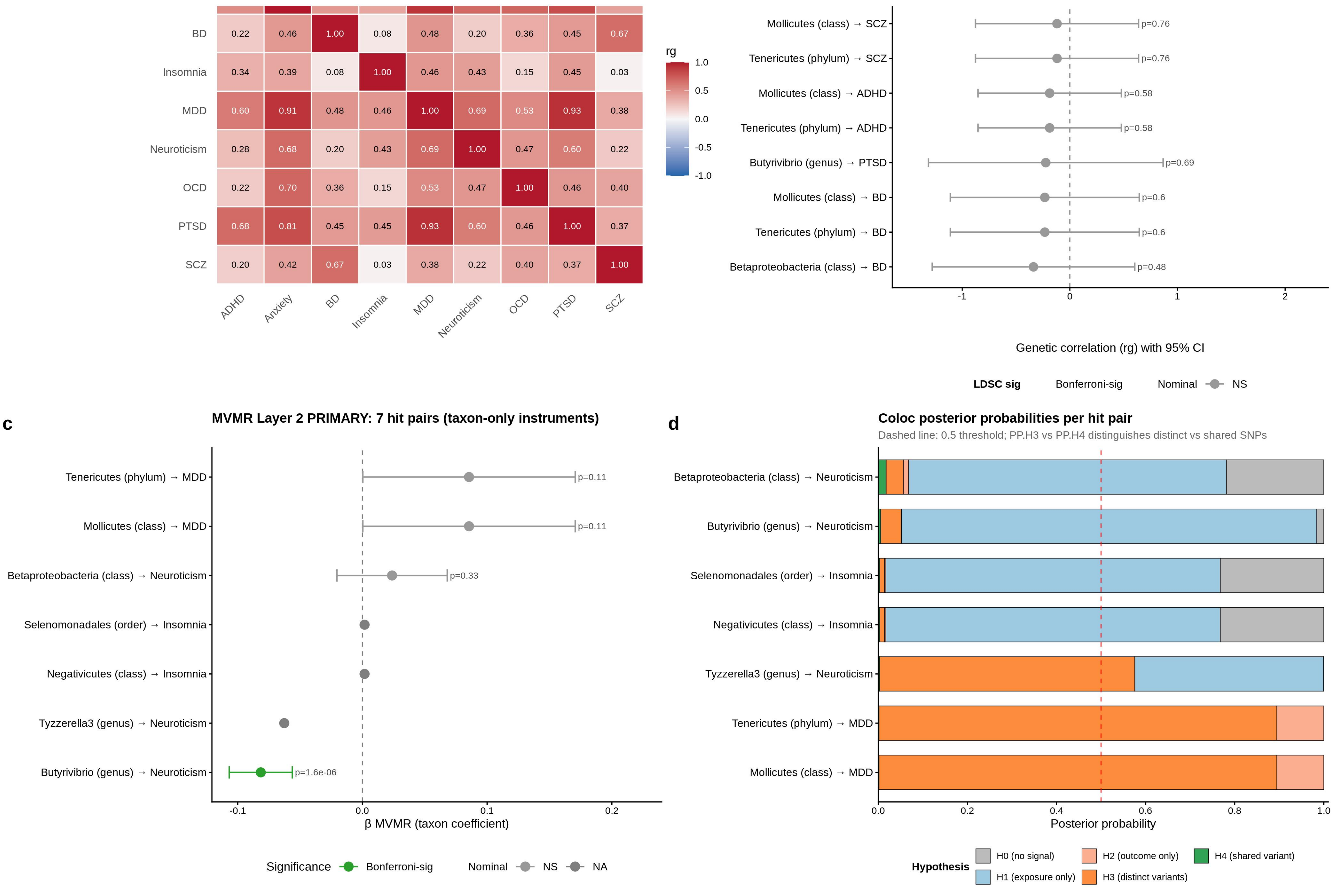
